# Prevalence and Clinical Implications of Heightened Plastic Chemical Exposure in Pediatric Patients Undergoing Cardiopulmonary Bypass

**DOI:** 10.1101/2023.05.02.23289379

**Authors:** Devon Guerrelli, Manan Desai, Youssef Semaan, Yasin Essa, David Zurakowski, Francesca I. Cendali, Julie A Reisz, Angelo D’Alessandro, Naomi C Luban, Nikki Gillum Posnack

## Abstract

**Importance:** Phthalate chemicals are used to manufacture disposable plastic medical products, including blood storage bags and components of cardiopulmonary bypass (CPB) circuits. During cardiac surgery, patients can be inadvertently exposed to phthalate chemicals that are released from these plastic products.

**Objective:** To quantify iatrogenic phthalate chemical exposure in pediatric patients undergoing cardiac surgery, and examine the link between phthalate exposure and post-operative outcomes.

**Design, Setting, and Participants:** The study cohort included 122 pediatric patients undergoing cardiac surgery at Children’s National Hospital. For each patient, a single plasma sample was collected pre-operatively and two additional samples were collected post-operatively upon return from the operating room (post-operative day 0) and the morning after surgery (post-operative day 1).

**Exposures:** Concentrations of di(2-ethylhexyl)phthalate (DEHP) and its metabolites were quantified using ultra high-pressure liquid chromatography coupled to mass spectrometry.

**Main Outcomes and Measures:** Plasma concentrations of phthalates, post-operative blood gas measurements, and post-operative complications.

**Results:** Study subjects were subdivided into three groups, according to surgical procedure: 1) cardiac surgery not requiring CPB support, 2) cardiac surgery requiring CPB with crystalloid prime, and 3) cardiac surgery requiring CPB with red blood cells (RBCs) to prime the circuit. Phthalate metabolites were detected in all patients, and postoperative phthalate levels were highest in patients undergoing CPB with RBC-based prime. Age-matched (<1 year) CPB patients with elevated phthalate exposure were more likely to experience post-operative complications, including arrhythmias, low cardiac output syndrome, and additional post-operative interventions. RBC washing was an effective strategy to reduce DEHP levels in CPB prime.

**Conclusions and Relevance:** Pediatric cardiac surgery patients are exposed to phthalate chemicals from plastic medical products, and the degree of exposure increases in the context of CPB with RBC-based prime. Additional studies are warranted to measure the direct effect of phthalates on patient health outcomes and investigate mitigation strategies to reduce exposure.

**Key Points:** *Question:* Is cardiac surgery with cardiopulmonary bypass a significant source of phthalate chemical exposure in pediatric patients?

*Findings:* In this study of 122 pediatric cardiac surgery patients, phthalate metabolites were quantified from blood samples before and after surgery. Phthalate concentrations were highest in patients undergoing cardiopulmonary bypass with red blood cell-based prime. Heightened phthalate exposure was associated with post-operative complications.

*Meaning:* Cardiopulmonary bypass is a significant source of phthalate chemical exposure, and patients with heightened exposure may be at greater risk for postoperative cardiovascular complications.

**Graphical Abstract:** Pediatric cardiac surgery patients undergoing cardiopulmonary bypass are exposed to phthalate plasticizers through contact with plastic medical devices manufactured with DEHP (indicated with blue outline), including blood product and crystalloid solution storage bags, tubing, cannulas, and reservoirs. The leached DEHP chemical (indicated with blue circles) can localize to the heart, where it can contribute to post-operative complications. DEHP: Di(2-ethylhexyl)phthalate, ET: endotracheal tube, FFP: fresh frozen plasma, RBCs: red blood cells.

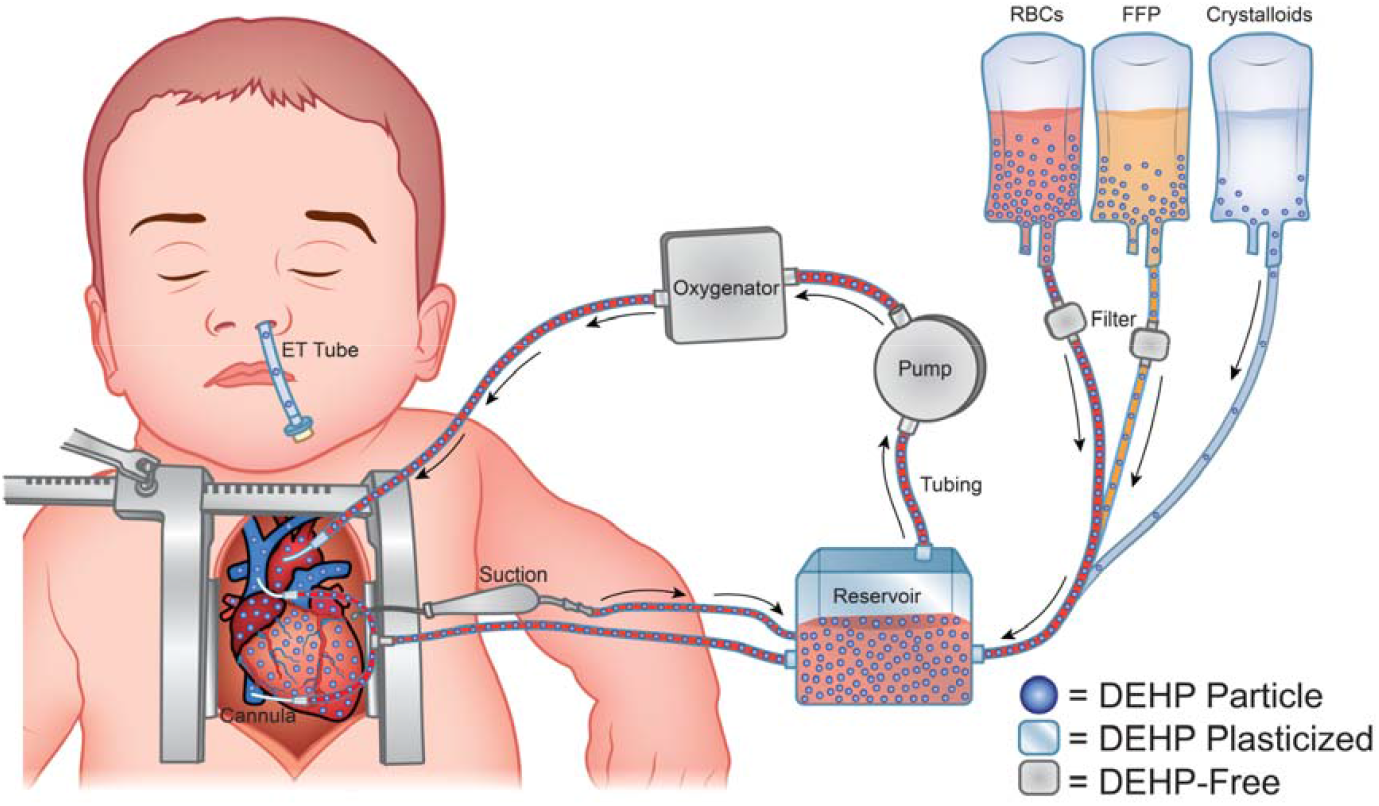

## Introduction

Phthalate plasticizers are chemical additives used to manufacture consumer and medical-grade plastics^1,2^. Di(2-ethylhexyl)phthalate (DEHP) is frequently employed in the production of polyvinyl chloride (PVC) plastics, wherein it embeds between PVC polymers to increase spacing and impart flexibility. DEHP is often added in large quantity, contributing up to 40-80% of the finished weight of medical-grade tubing and blood storage bags^3,4^. Since phthalates are not covalently bound to PVC, these chemicals are highly susceptible to leaching, particularly in the presence of lipophilic solutions^5–7^. Concerns over the safety of inadvertent phthalate exposure through contact with medical plastics have existed since the 1970s^4,8–10^.

Phthalate exposure is particularly concerning for neonatal and pediatric intensive care unit (NICU, PICU) patients who are treated using multiple plastic devices^11–15^. Studies have reported increased urinary phthalate concentrations in NICU patients^16–20^, which can exceed safety thresholds for reproductive and hepatic toxicity by >1,000-fold^21^. Notably, phthalate chemicals have been found to localize to the heart tissue of infants requiring umbilical catheterization and/or transfusion^22,23^. Further, in vitro studies have reported a direct causation between phthalate exposure and cardiotoxicity, including a negative inotropic effect on human atrial trabecula^24^. Rodent studies have reported similar cardiodepressive effects after phthalate exposure, resulting in hypotension, bradycardia, cardiac arrest, and increased arrhythmia incidence^25–27^.

This study aimed to quantify phthalate chemical exposure, including DEHP and its metabolites, in pediatric patients undergoing cardiac surgery with/without cardiopulmonary bypass (CPB). We also aimed to provide information on the timing of phthalate clearance in the post-operative period. Finally, we assessed possible correlations between higher phthalate exposures and post-operative complications.

## Methods

### Study Population

Experiments were conducted in accordance with human research guidelines; the study was approved by the Children’s National Hospital Institutional Review Board (Pro00009620). For children old enough to comprehend the study, the patient’s assent was obtained in addition to parental permission. The study population was divided into three groups **(Figure 1, Supplemental Table 1)**. *<u>Group 1 “No CPB”</u>* served as a surgical control and included patients undergoing a procedure via thoracotomy without CPB (n=11). These surgeries were mostly coarctation repairs or placement of pulmonary artery bands, wherein the only identified source of phthalate exposure was the endotracheal tube. CPB patients were subdivided based on the bypass prime composition, since DEHP leaching is prevalent in RBC units but limited in crystalloid solutions devoid of lipids^7^. *<u>Group 2 “Crystalloid prime”</u>* included CPB patients (n=21) with a crystalloid based prime, with (n=12) or without (n=9) fresh frozen plasma (FFP) supplementation. *<u>Group 3 “RBC prime”</u>* included CPB patients with an RBC-based prime (n=90). Patients received either unwashed RBCs (n=33) or RBCs washed in a Sorin XTRA cell saver (n=57).

**Figure 1.**
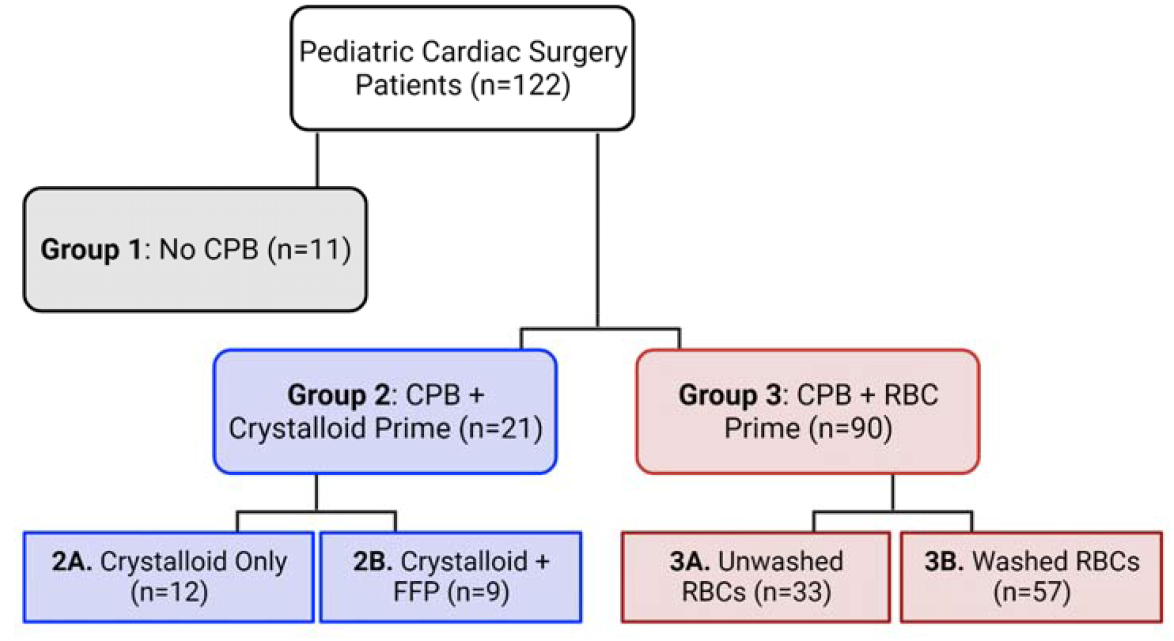
Patient Groups. Pediatric cardiac surgery patients (n=122) were enrolled in this study. Patients were subdivided into those undergoing cardiac surgery without CPB (Group 1, n=11) or with CPB (Group 2, 3). CPB patients were further subdivided based on the CPB prime. Patients receiving a crystalloid prime with or without the addition of FFP were grouped (Group 2, n=21), and patients receiving RBC-based prime were grouped (Group 3, n=90). In the latter, patients may receive unwashed RBCs (n=33) or washed RBCs (n=57), at the discretion of the perfusionist.

### Cardiopulmonary Bypass

Blood products used at our institution for CPB are less than 8 days old and x-ray irradiated at time of issue. For patients weighing <5 kg, RBCs were washed to remove extracellular potassium and lactate which accumulate during storage and following irradiation. For patients weighing 5-15 kg, unwashed RBCs were used, since it is assumed that lactate is more easily cleared in these patients. For patients weighing >15 kg, RBCs were not used in the prime, as long as the estimated hematocrit was deemed acceptable by the perfusionist. However, RBCs would still be used if significant hemodilution was expected. Further, RBCs were transfused if significant bleeding occurred. In a subset of patients, methylprednisolone was administered to reduce CPB-induced inflammation, which can be more significant in younger, smaller patients^28^. Neonates (<1 month old) received methylprednisolone on the morning of their surgery and 10 mg/kg methylprednisolone was added to the CPB prime.

### Patient & RBC Unit Sample Collection

Plasma samples were obtained from specimens collected for routine clinical labs at Children’s National Hospital. A pre-operative sample (pre-op) was predominately collected the day before or the morning of surgery. Post-operative samples were drawn upon arrival to the cardiac intensive care unit (post-operative day 0, “POD0”) and ∼4 am on the morning after surgery (post-operative day 1, “POD1”). To evaluate whether washing RBC units altered phthalate levels, an in vitro study was performed (n=4) wherein samples were collected before and after cell salvage with a cell saver. Samples were frozen and shipped to the University of Colorado for quantification of DEHP and metabolites (MEHP: mono(2-ethylhexyl)phthalate, MEHHP: mono(2-ethyl-5-hydroxyhexyl)phthalate, MECPP: mono(2-ethyl-5-carboxypentyl)phthalate; see **Supplemental Methods**).

### Ascertainment of Post-Operative Outcomes

Clinical data was retrospectively collected from medical records (see **Supplemental Methods**); patient characteristics, laboratory values, post-operative complications, within the first 48 hours after surgery were documented. To remove age as a confounding factor, only patients <1 year old were included in post-operative complication and intervention analysis.

### Statistical Analysis

Results are reported as the mean ± standard deviation. Normality was assessed using a Shapiro-Wilk test and variance was assessed using an F-test (two groups) or Barlett’s test (three groups). <u>For repeated measurements</u> (e.g., pre- and post-operative phthalate levels in the same patient), normally distributed data were analyzed via one-way ANOVA without or with Geisser-Greenhouse correction (unequal variance). Nonparametric data were analyzed via a Friedman test. <u>For independent measurements</u> (e.g., phthalate levels between patients, blood gas), normally distributed data were analyzed via one-way ANOVA without or with Welch’s correction (unequal variance). Nonparametric data were analyzed via Kruskal-Wallis ANOVA. A false discovery rate (FDR, 0.1 cutoff) was used to correct for multiple comparisons^29–31^. Two-tailed Student’s t-test was used to compare phthalate levels and post-operative complications without or with Welch’s correction (unequal variance). Spearman correlations were used to measure the association between independent surgical variables (e.g., CPB time, index prime volume) and total phthalate exposure. A p-value <0.05 was considered statistically significant.

## Results

### Change in Pre- and Post-operative Phthalate Levels in Cardiac Surgery Patients

Previous studies reported routine phthalate exposure in the general population through daily contact with plastic consumer products^32,33^. In agreement, we detected DEHP and its metabolites in all pre-operative blood samples, regardless of inpatient or outpatient status **(Figure 2, Supplemental Table 2)**. The pre-operative ‘phthalate sum’ (DEHP and its metabolites) was comparable across groups (No CPB: 1.37±0.73 μM, Crystalloid prime: 1.42±0.65 μM, Crystalloid prime+FFP: 1.59±1.01 μM, RBC-based prime: 1.96±2.56 μM, p>0.05). The ‘No CPB’ group had a minimal 0.8% increase in their phthalate sum after surgery (POD0), compared to their pre-operative level **(Figure 2E)**. Patients receiving crystalloid prime, crystalloid prime+FFP, or RBC-based prime had a 93.9%, 176%, or 481% increase in their phthalate sum concentration after surgery, respectively, compared to their pre-operative levels **(Figure 2 J,O,T)**. Patients receiving RBC-based prime had an increase in every metabolite measured: 193% increase in DEHP (Pre-op: 1.29±1.51, POD0: 3.79±2.24 μM), 1064% increase in MEHP (Pre-op: 0.44±1.32, POD0: 5.12±4.37 μM), 1221% increase in MECPP (Pre-op: 0.14±0.28, POD0: 1.85±1.46 μM), and 611% increase in MEHHP (Pre-op: 0.09±0.25, POD0: 0.64±0.43 μM; **Figure 2P-S**).

**Figure 2:**
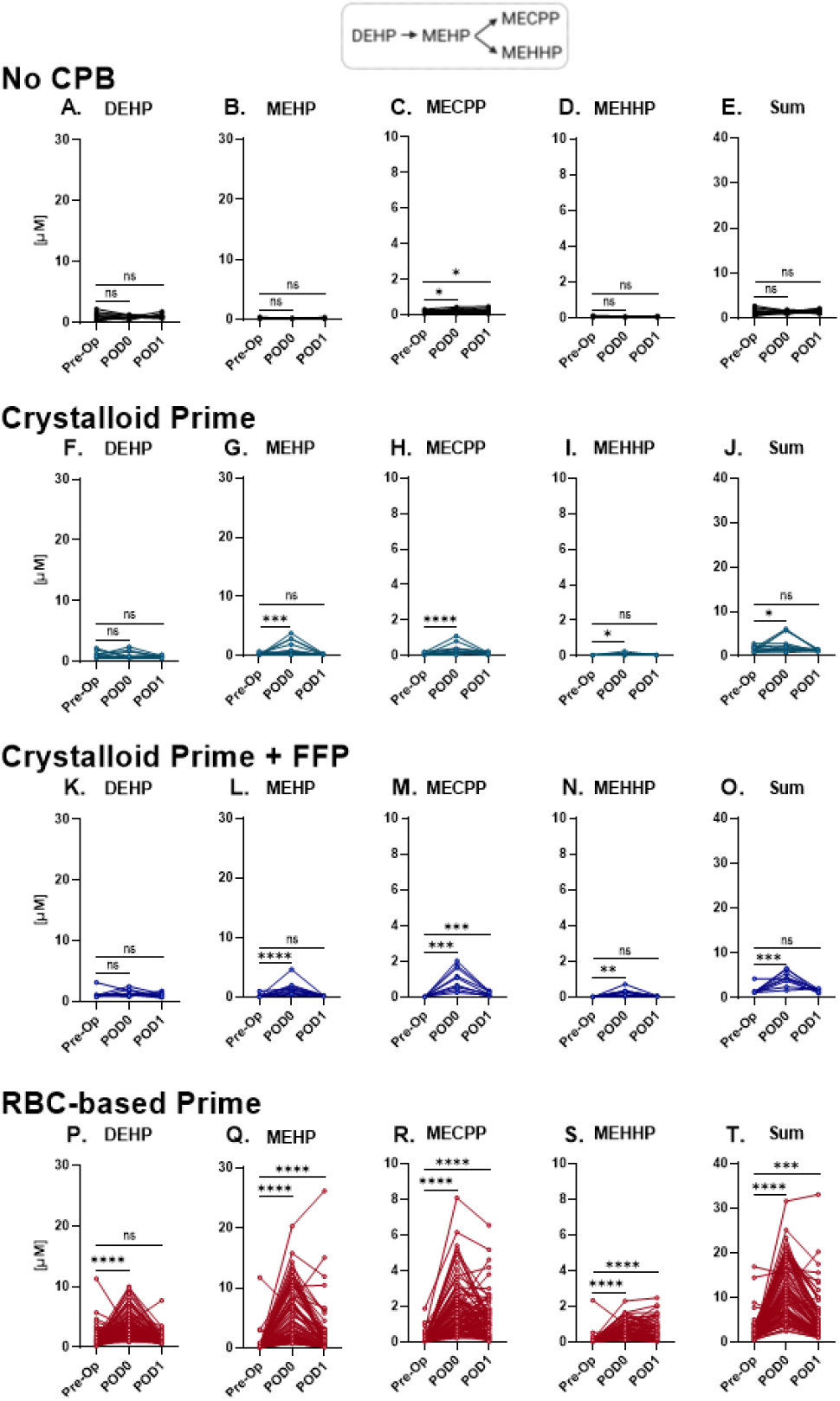
Phthalate chemical concentrations increase in patient blood samples afte cardiac surgery. **A-E)** Patients undergoing cardiac surgery without cardiopulmonary bypass (No CPB). **B)** Patients undergoing CPB without the use of blood products. **F-J)** Patients undergoing CPB with fresh frozen plasma (FFP). **K-O)** Patients undergoing CPB with red blood cells (RBC) and FFP. Levels of DEHP, its metabolites (MEHP, MECPP, MEHHP), and the sum of all phthalate concentrations are reported. Matched values for each individual patient are connected by a line. Phthalate levels were measured at pre-operative visit (Pre-op); post-operative day 0 (POD0); and post-operative day 1 (POD1). *Repeated measurements: normally distributed data analyzed via repeat measures ANOVA (equal variance) with Geisser-Greenhouse correction (unequal variance). Nonparametric data analyzed via Friedman test. FDR (0*.*1 cutoff) was used to correct for multiple comparisons; statistical significance denoted by *p<0*.*05, **p<0*.*01, ***p<0*.*005, ****p<0*.*0001, ns=not significant*

By POD1, phthalate sum had declined to ±15% of the pre-operative level in the ‘No CPB’ and ‘Crystalloid prime±FFP’ groups. However, there was a persistent elevation in MECPP, a secondary metabolite of DEHP, at this later time point relative to the pre-operative measurements (No CPB: +45.1%, Crystalloid+FFP prime: +307.3%; **Figure 2 C,H,M**). In the RBC-based prime group, the phthalate sum remained 146.6% higher at POD1 compared to pre-operative baseline **(Figure 2T)**. At POD1, DEHP levels returned to baseline, but primary and secondary metabolites remained elevated (MEHP: +309.2%, MECPP: +787.1%, MEHHP: +377.2%; **Figure 2P-S**). Since phthalate concentrations were comparable between CPB patients receiving either crystalloid-based prime or crystalloid-based prime+FFP, these patients were combined for further analyses.

### Phthalate Exposure Varies by Surgical Intervention and Age

Previous studies have reported increased DEHP exposure in NICU patients receiving multiple medical interventions^14,19,20,34^. Transfused patients can also be exposed to phthalates that leach and accumulate in blood bags during storage^35,36^. We found that patients undergoing cardiac surgery without CPB had the lowest phthalate levels after surgery (1.38±0.32 μM phthalate sum; **Figure 3A-E**). Phthalate levels were higher in patients undergoing CPB with crystalloid prime±FFP (3.45±1.97 μM phthalate sum), which may be attributed to plastic materials in the CPB circuit. At POD0, MEHP was the most abundant metabolite detected in this group (1.42±1.23 μM MEHP). Patients undergoing CPB with RBC-based prime had the highest phthalate levels (11.40±6.19 μM phthalate sum). MEHP was also the most abundant metabolite detected in this group at POD0 (5.12±4.37 μM MEHP), indicating that RBCs are also a major source of phthalate exposure. Although, direct conclusions cannot be made due to significant differences in patient demographics between groups (e.g., age of patients in Group 3: 194.5±410 days versus Group 2: 2563±1785 days, p<0.0001). We also sought to understand how quickly phthalates are cleared from patients’ blood. Pharmacokinetic studies indicate that oral DEHP exposure results in rapid metabolism to MEHP (∼5-hour half-life), with a subsequent increase in secondary metabolites including MEHHP and MECPP (∼10-15 hour half-life^37,38^). All DEHP metabolites remained elevated at POD1 in patients undergoing CPB with RBC-based prime (**Figure 3, Supplemental Table 2**).

**Figure 3:**
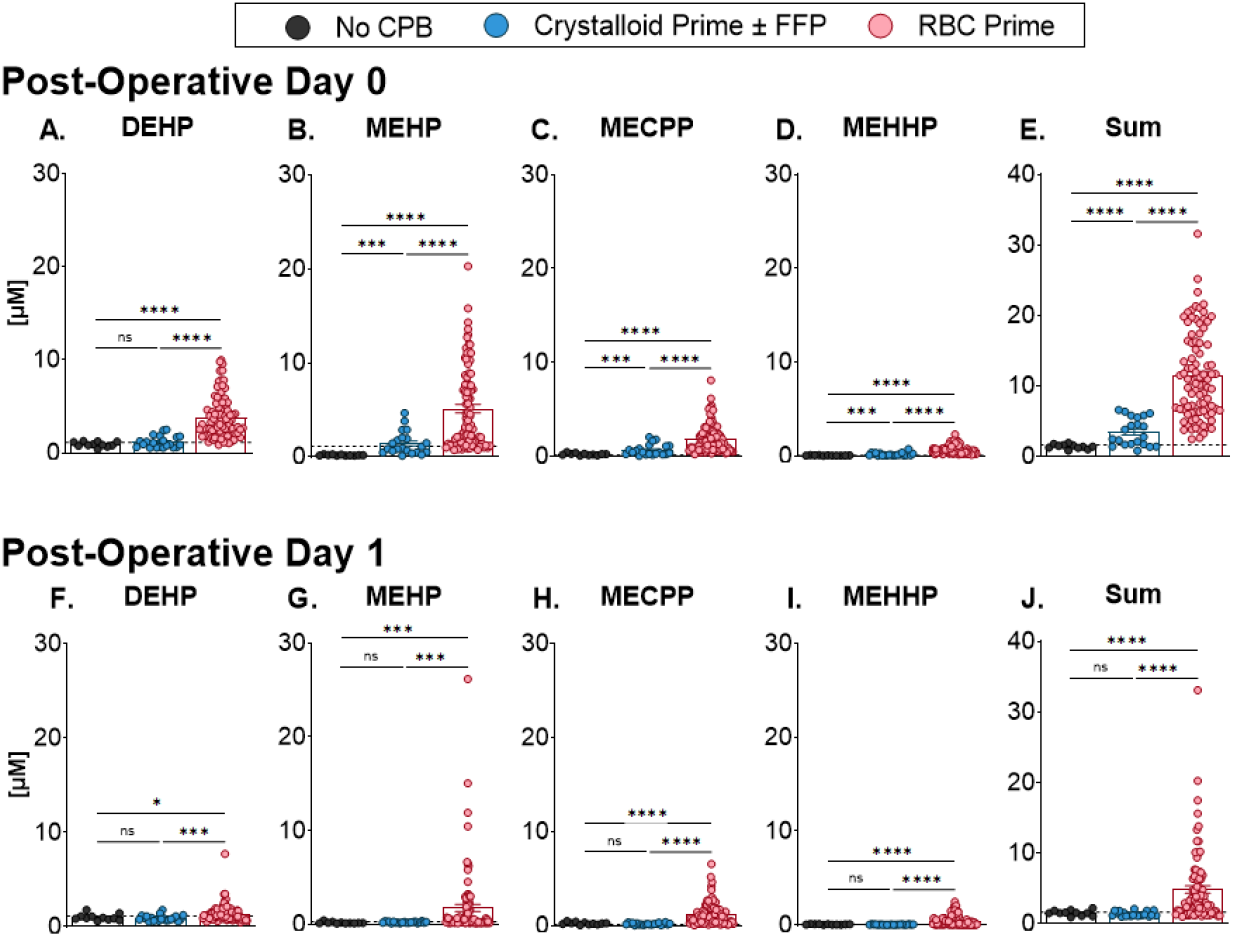
Higher phthalate chemical concentrations are detected in patients requiring CPB with RBC-based prime. **A-E)** DEHP, MEHP, MECPP, MEHHP, and total phthalate concentration (sum) in blood samples measured at post-operative day 0, and **F-J)** post-operative day 1. For comparison, the mean preoperative value (all three groups combined) is indicated by a dashed line. *Independent measurements analyzed via one-way ANOVA with Welch’s correction (unequal variance). FDR (0*.*1 cutoff) was used to correct for multiple comparisons; statistical significance denoted by *p<0*.*05, **p<0*.*01, ***p<0*.*005, ****p<0*.*0001, ns=not significant*.

Next, we explored variations in the CPB procedure that could contribute to phthalate exposure, since phthalate sum levels ranged broadly in CPB patients (0.77-31.57 μM, POD0). For example, in young patients, underdeveloped glucuronidation and/or reduced renal function could diminish their ability to metabolize and excrete phthalates^39^. We found a significant correlation between age and phthalate concentration (r=-0.81, p<0.001), wherein infants <1 year old had increased phthalate levels (12.34±6.0 μM) compared to patients >1 year old (3.87±2.05 μM phthalate sum; **Figure 4A**). Published *in vitro* experiments suggest that phthalate leaching from tubing is influenced by surface area, flow rate, temperature, perfusion time and/or storage duration^7,36,40–43^. Since CPB flow rate is calculated by multiplying the body surface area by cardiac index (2.4 L/min/m^2^)^44^, flow rate in younger patients is inherently tied to age and body surface area. Therefore, we measured the indexed flow rate (flow rate/body surface area), which showed no correlation to phthalate exposure (**Figure 4B**). Absolute prime volume is also influenced by age, as younger patients receive a smaller CPB prime volume. We measured the prime volume indexed by body surface area and found a positive correlation with phthalate levels (r=0.67, p<0.0001; **Figure 4C**).

**Figure 4:**
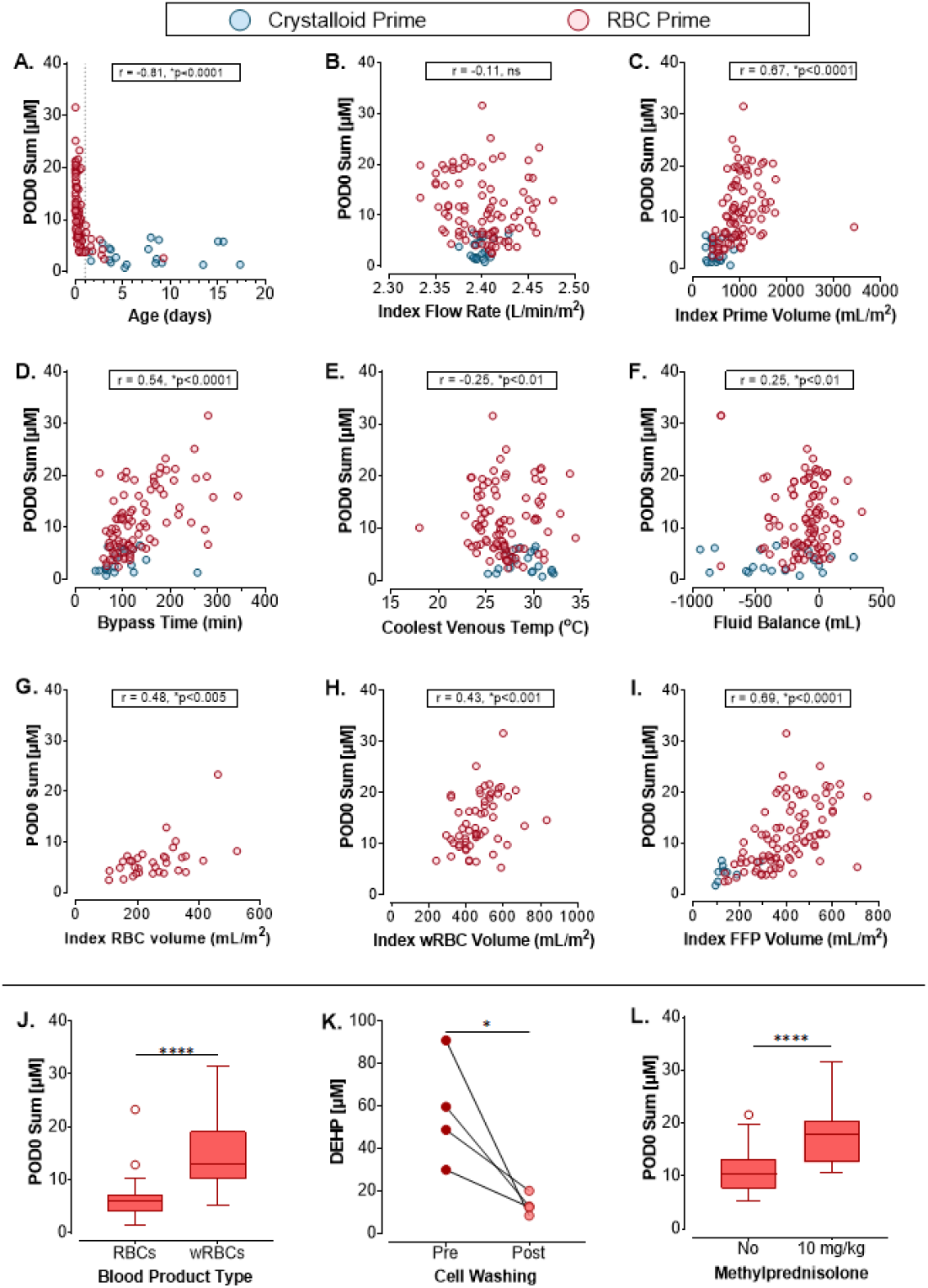
Surgical variables that may contribute to heightened phthalate exposure. **A)** Phthalate levels were elevated in patients <1 year old. **B)** Phthalate levels did not correlate with index flow rate. **C)** Phthalate levels increased with prime volume. **D-F)** Phthalate levels increased with CPB time, and there was a weak correlation with venous temperature and fluid balance. **G-I)** Phthalate levels did not correlate with unwashed RBC blood volume or FFP volume, but there was a correlation with washed RBC volume. **J)** Patients <5 kg received washed RBCs (wRBC) and had higher phthalate blood levels. **K)** Cell washing procedure alone did not increase phthalate concentrations in an *in vitro* study. **L)** Heightened phthalate levels in patients receiving washed RBC, may instead be influenced by administration of steroids (10 mg/kg methylprednisolone) that alters metabolism. POD0 Sum = total phthalate concentration measured on post-operative day 0. *(A-I) Spearman correlation indicated for each variable, with denoted p-value. (J,L) Two-tailed student’s t-test. (K) Two-tailed repeat measures t-test. *p<0*.*05, ****p<0*.*0001*

DEHP leaching from tubing is also influenced by the temperature of the solution and duration of contact ^40,43^, with an approximate leaching rate of 3.12 grams/L/hour using a heart-lung machine^45^. In agreement, we found that longer bypass time was associated with increased phthalate exposure (r=0.54, p<0.0001; **Figure 4D**). We anticipated that therapeutic hypothermia during CPB might reduce phthalate exposure, but detected only a modest association (r=-0.25, p<0.01; **Figure 4E**). This is likely due, in part, to patients who undergo longer bypass times also undergoing therapeutic hypothermia. Finally, we found a modest correlation between patient fluid balance and post-surgical phthalate levels (r=0.25, p<0.01; **Figure 4F**).

Finally, we assessed whether phthalate exposure correlates with blood product usage as 91.9% of CPB patients received FFP, 29.7% received unwashed RBCs, and 51.4% received washed RBCs. Indexed volumes of unwashed RBCs (r=0.48, p<0.005), washed RBCs (r=0.43, p<0.001), and FFP (r=0.69, p<0.0001) were positively correlated with POD0 phthalate levels **(Figure 4G,H)**. Unfortunately, we do not know which patients received blood products stored in DEHP vs DEHP-free bags (spot checking estimated >97% of RBC units and >70% of FFP units were manufactured with DEHP). Controlling for age (<1 year old), we compared phthalate levels in CPB patients who received either unwashed or washed RBCs and found that the latter had a 2.3-fold increase in phthalate levels (unwashed: 6.15±3.7 μM, washed: 14.3±5.4 μM phthalate sum; **Figure 4J**). This result was unexpected, as cell washing removes extracellular contaminants from RBC units^46^. We hypothesized that increased contact with plastic materials during cell washing might increase phthalate leaching. However, our *in vitro* study showed that cell washing reduced DEHP levels in RBC units by 76.6% (pre-washing: 57.3±25.5 M, post-washing: 13.4±4.9 μM DEHP; **Figure 4K**). We speculate that higher phthalate levels in patients receiving washed RBCs might be attributed to perioperative administration of steroids, as 55.7% of patients receiving washed RBCs were also administered 10 mg/kg methylprednisolone. These patients had a 55.3% increase in phthalate levels compared to weight-matched controls (no steroids: 11.2±4.4 μM, 10 mg/kg methylprednisolone: 17.4±5.0 μM), suggesting that steroid effects on lipase activity may be influencing phthalate metabolism in this group^47,48^.

### Phthalate Exposure is Associated with Altered Electrolyte Balance

We investigated trends between phthalate exposure and post-operative blood gas parameters in CPB patients, as experimental work suggests that DEHP alters serum biochemistry^49^. We measured higher phthalate levels in CPB patients who presented with hypokalemia (low K^+^: 12.5±6.7 μM; normal K^+^: 8.3±5.8 μM phthalate sum), hypernatremia (high Na^+^: 11.7±6.7 μM; normal Na^+^: 7.8±5.5 μM phthalate sum), and a higher anion gap (high anion gap: 12.9±6.7 μM; normal anion gap: 7.0±4.6 μM; see **Supplemental Figure 1**). Elevated fasting glucose levels have also been reported in individuals with higher daily phthalate exposure ^50^. Similarly, we found elevated phthalate levels in patients with hyperglycemia requiring intervention (high glucose: 10.8±6.9 μM; normal glucose: 8.1±5.1 μM phthalate sum). Phthalate exposure was inversely associated with hypocalcemia (low calcium: 3.3±2.2 μM; normal calcium: 9.8±6.6 μM phthalate sum), lower lactate levels (low lactate: 5.4±3.9 μM; normal lactate: 10.5±5.8 μM phthalate sum), and acidosis (low pH: 3.3±1.6 μM; normal pH: 11.07±6.5 μM phthalate sum; see **Supplemental Figure 1**).

### Heightened Phthalate Exposure is Associated with Post-Operative Complications

In experimental models, phthalate exposure precipitates cardiac arrhythmias^26,51^. Accordingly, we investigated associations between phthalate exposure and 48-hour postoperative outcomes in age-matched (<1 year) CPB patients. Phthalate levels were lower in patients without postoperative complications (6.1±2.6 μM phthalate sum), and higher in patients experiencing cardiac arrhythmias or mechanical instability, including junctional ectopic tachycardia (16.4±7.0 μM phthalate sum), supraventricular tachycardias (13.5±6.5 μM phthalate sum), ventricular tachycardia/fibrillation (20.6±2.6 μM phthalate sum), and heart block (13.0±6.4 μM phthalate sum, **Figure 5A-E**). Increased phthalate levels were also associated with low cardiac output syndrome and/or cardiogenic shock (14.7±6.3 μM phthalate sum), cardiac arrest (12.0±6.8 μM phthalate sum), hypotension (13.3±5.3 μM phthalate sum), and blood pressure liability (18.0±4.7 μM phthalate sum), wherein a patient’s blood pressure is unstable (**Figure 5G-K**). We did not find a link between phthalate exposure and hypertension (**Figure 5J**). Studies suggest that phthalate exposure is associated with kidney injury^52,53^, and we found a positive association between phthalate exposure (16.0±9.5 μM phthalate sum) and acute kidney injury or renal insufficiency (**Figure 5L**). It has also been documented that phthalates alter cellular metabolism^54–56^, and we observed a positive association between phthalate exposure and lactic acidosis or hyperglycemia requiring intervention (15.5±6.1 μM and 21.3±6.7 μM phthalate sum, respectively; **Figure 5M-Q**). Finally, we noted that patients with elevated phthalate levels were more likely to require interventions after surgery, including extracorporeal membrane oxygenation (ECMO) and transfusion (13.6±6.5 μM and 12.9±5.4 μM phthalate sum, respectively; **Figure 5Q,R**).

**Figure 5:**
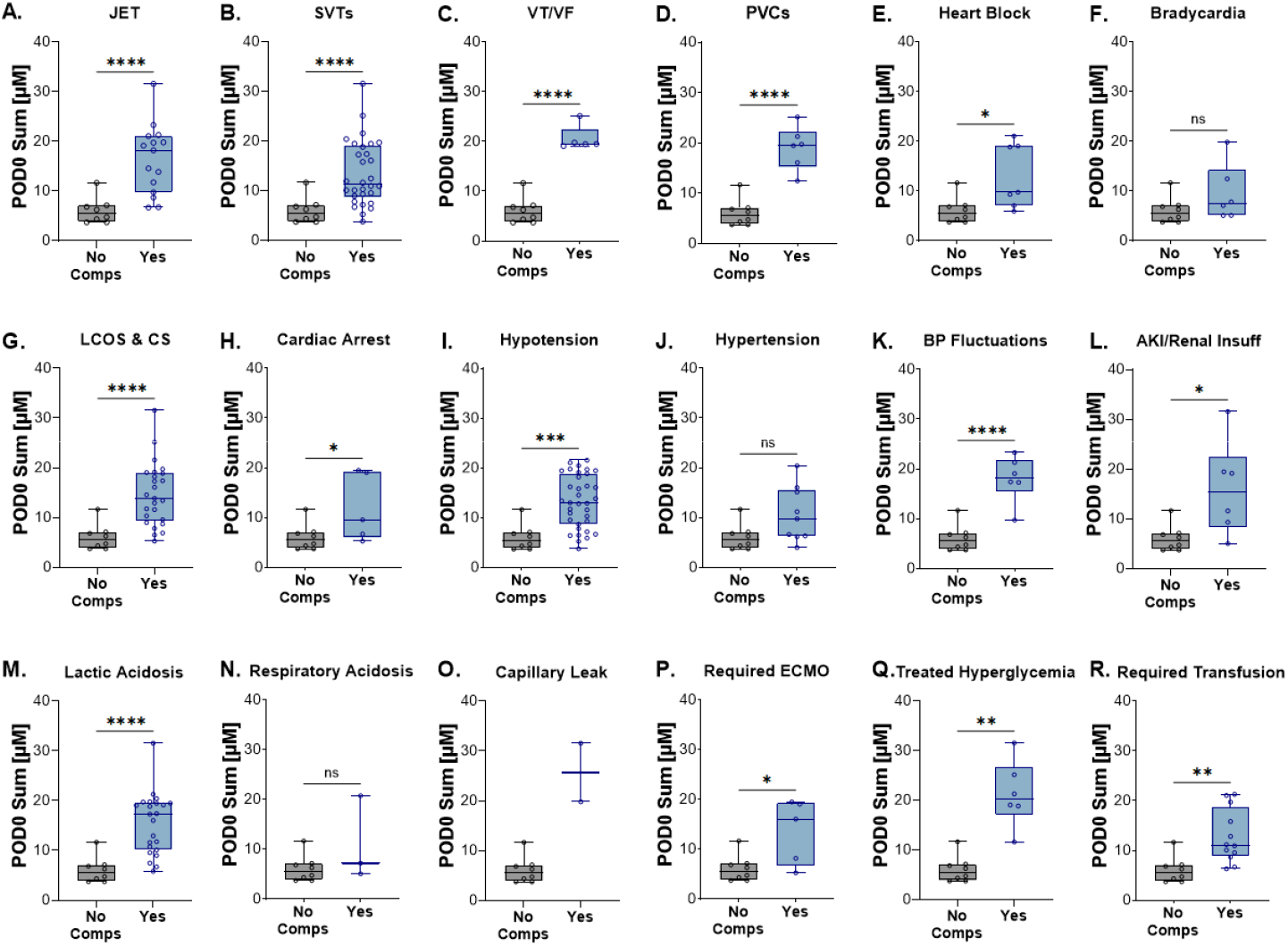
Association between phthalate levels and 48-hour post-operative outcomes in cardiac surgery patients <1 year old. Associations between elevated plasma phthalate levels on post-operative day 0 and post-operative complications including: **A)** junctional ectopic tachycardia (JET), **B)** supraventricular tachycardias (SVTs), **C)** ventricular tachycardia or fibrillation (VT/VF), **D)** premature ventricular contractions (PVCs), **E)** heart block, **F)** bradycardia, **G)** low cardiac output syndrome (LCOS) and/or cardiogenic shock (CS), **H)** cardiac arrest, **I)** hypotension, **J)** hypertension, **K)** blood pressure (BP) fluctuations, **L)** acute kidney injury (AKI) or renal insufficiency, **M)** lactic acidosis, **N)** respiratory acidosis, **O)** capillary leak. Association between elevated plasma phthalate levels and additional post-operative treatment, including **P)** extracorporeal membrane oxygenation (ECMO), **Q)** intervention for hyperglycemia, **R)** additional transfusion. *Statistical analysis by two-tailed student’s t-test (equal variance), with Welch’s correction (unequal variance). *p<0*.*05, **p<0*.*01, ***p<0*.*005, ****p<0*.*0001*

## Discussion

We quantified phthalate plasticizer exposure in pediatric patients undergoing cardiac surgery. DEHP and its metabolites were detected in all patient samples, but higher phthalate levels were detected in patients undergoing CPB with RBC-based prime and phthalate levels remained elevated until the day after surgery (later time points were not investigated). Higher phthalate levels were associated with younger patient age, longer bypass times, greater indexed prime volume, fluid balance, and RBC-based CPB prime. Our *in vitro* study showed that washing RBCs was an effective strategy toward reducing phthalate levels in CPB prime. However, unexpectedly, we detected elevated phthalate levels in patients <5kg who received washed RBCs – which may be attributed to the immature metabolic capacity of these younger patients and/or the co-administration of methylprednisolone which alters lipase activity and phthalate metabolism^48,57^. We also identified associations between phthalate exposure and post-operative complications, including cardiac electrical instabilities, impaired contractile function, blood pressure disturbances, and an imbalance in blood gas measurements.

In agreement with prior studies, our work identifies cardiac surgery and circulatory support as a clinical source of phthalate exposure. Barry et al. reported elevated DEHP blood levels in infants undergoing CPB, ranging from 1.1-5.6 μg/mL (n=7)^58^. Gaynor et al. also found increased urinary DEHP metabolite levels in infants after cardiac surgery (n=18)^59^. Eckert et al. reported an increased blood phthalate levels in infants undergoing CPB (n=21), despite the use of DEHP-free tubing. Specifically, phthalate exposure was attributed to DEHP accumulation in stored RBC (6.3-22.2 μg/mL DEHP) and FFP units (0.77-2.03 μg/mL DEHP)^60^. Finally, Karle et al. reported elevated plasma phthalate concentrations in infants receiving ECMO (0-24.2 μg/mL DEHP, n=18)^42^. Phthalates have been shown to localize to the heart tissue of infants who received umbilical catheters and/or blood transfusions prior to death^22,23^. Our work expands on these reports, benefitting from a larger study enrollment across a wider age range, which can help to better understand phthalate exposure and metabolism in pediatric patients. Moreover, our study includes a control group of surgery patients undergoing a thoracotomy without CPB, which helps to distinguish exposures from surgery alone versus plastic tubing or blood product usage. Finally, we show that cell washing removes DEHP from fresh RBC units, which builds upon the work by Münch et al. demonstrating that washing decreases DEHP and MEHP levels by 18-96% in 36-56 day old blood units^61^.

Clinical studies linking phthalate exposure to patient outcomes are sparse, but experimental work has demonstrated that phthalates exert cardiotoxic effects^2,62,63^. DEHP exposure halted the spontaneous beating activity of chick cardiomyocytes^8^ and slowed heart rate, atrioventricular conduction, and impaired contractile activity in rodent models^26,27,64^. Further, MEHP infusion can precipitate hypotension and cardiac arrest in rodents^51^. These experimental findings support the postoperative complications observed in this study, wherein CPB patients <1 year with heightened phthalate exposure were more likely to experience cardiac arrest, post-operative arrhythmias, and reduced contractile function. Taken together, it is plausible that a combination of risk factors (young age, longer bypass duration, increased phthalate exposure) collectively contribute to postoperative complications. Notably, the Food and Drug administration issued a recommendation to minimize phthalate exposure in neonatal boys due to reproductive toxicity concerns^65^. This prompted NICUs to adopt alternative plastic products^66,67^; yet the same efforts have not been applied to pediatric cardiac intensive care units or cardiothoracic surgery. Future studies should investigate mitigation strategies to reduce phthalate exposure, including adopting alternative tubing materials that are less prone to leaching, using DEHP-free blood bags, implementing cell washing to remove phthalates, and/or decreasing CPB prime volume.

## Study Limitations

Our study population is heterogeneous, as the enrolled patients differ in the complexity of their congenital heart defects and surgical repair. Therefore, we cannot prove causation between phthalate exposure and post-operative complications, and additional work is needed to evaluate these associations further. Our study used blood samples that were drawn in accordance with clinical care, therefore, we could not standardize the exact time of sample collection. Peak blood phthalate concentrations are likely to occur during or immediately after surgery, yet the earliest time point we measured (POD0) coincided with arrival to the cardiac intensive care unit. POD1 samples were collected at approximately 4 AM the morning after surgery, which could lead to slight variations in the length of time between end of surgery and sample collection. Finally, post-operative outcomes were assessed retrospectively and were limited to the thoroughness of daily progress notes.

## Data Availability

The participants of this study did not give written consent for their data to be shared publicly; data to support the findings of this study are available on request from the corresponding author.

## Abbreviations

CPB: Cardiopulmonary bypass
DEHP: Di(2-ethyhexyl)phthalate
FDR: False discovery rate
FFP: Fresh frozen plasma
MECPP: Mono(2-ethyl-5-carboxypentyl)phthalate
MEHHP: Mono(2-ethyl-5-hydroxyhexyl)phthalate
MEHP: Mono(2-ethylhexyl)phthalate
NICU: Neonatal Intensive Care Unit
PICU: Pediatric Intensive Care Unit
POD0: Post-operative day 0
POD1: Post-operative day 1
Pre-op: Pre-operative
RBCs: Red blood cells

## Acknowledgements

The authors acknowledge Daniel Stephenson, PhD for assistance with metabolomics analysis and Bryan Siegel, MD for contributions to the study concept. We additionally acknowledge the Pathology & Laboratory Medicine Research Core at Children’s National Hospital for assistance with biospecimen retrieval including Meghan Delaney DO, Willian Suslovic, Casey Yarbrough, Aszia Burrell, and Eric Freeman.

