## Supplemental File for "Prevalence and Clinical Implications of Heightened Plastic Chemical Exposure in Pediatric Patients Undergoing Cardiopulmonary Bypass"

**Supplemental Methods**

Phthalate Chemical Extraction and Quantitation

Phthalate chemical concentrations were measured in both patient plasma and in samples collected from RBC (red blood cell) and wRBC (washed red blood cell) units. Briefly, aliquoted samples were shipped on dry ice to the University of Colorado School of Medicine Metabolomics Core. Samples were thawed on ice and phthalates were extracted from a 10 μL aliquot via vigorous vortexing for 30 min at 4°C in the presence of 5:3:2 methanol:acetonitrile:water (240 μL) containing 1 μM of each of the following stable isotope labeled phthalate standards (Cambridge Isotope Laboratories): DEHP ring-1,2-^13^C_2_, dicarboxyl-^13^C_2_; MEHP ring-1,2-^13^C_2_, dicarboxyl-^13^C_2_; MECPP ^13^C_4_; MEHHP ^13^C_4_ (DEHP: di(2-ethyhexyl)phthalate, MEHP: mono(2-ethylhexyl)phthalate, MECPP: mono(2-ethyl-5-carboxypentyl)phthalate, MEHHP: mono(2-ethyl-5-hydroxyhexyl)phthalate). Extraction supernatants were clarified via centrifugation at 18,000 rpm for 10 min at 4°C and analyzed immediately by ultra high-pressure liquid chromatography coupled to mass spectrometry (UHPLC-MS). Sample injection volume was 20 μL. Samples were analyzed on a Thermo Vanquish UHPLC coupled to a Thermo Q Exactive high resolution mass spectrometer. The LC was equipped with a Phenomenex Kinetex C18 column (2.1 x 150 mm, 1.7 μm) held at 45 °C (gradient information below). The mass spectrometer scanned in MS1 mode in the range of 65 to 975 m/z at a resolution of 70,000. Instrument-generated .raw files were converted to .mzXML format via RawConverter and peaks for phthalates and accompanying standards were extracted and integrated using Maven (Princeton University). Absolute concentrations were obtained using the following equation where dilution factor (DF) is 25:

[Phthalate] = (Peak Area Phthalate) / (Peak Area Standard) * [Standard] * DF

DEHP was quantified using a 5 min C18 gradient at a flow rate of 450 μL per min as previously described^40^ with a single change – eluant was introduced to the MS using atmospheric pressure chemical ionization (APCI) in positive mode. APCI was performed using 10 sheath gas, 20 aux gas (both N_2_), and 350 °C vaporizer temperature. The calculations of DEHP concentrations utilized the average peak area of the stable isotope labeled DEHP standard across all samples within the biological group.

Monoesterified phthalates (i.e., MEHP, MECPP, MEHHP) were quantified using a 5 min C18 gradient with phase A of 5 mM ammonium acetate, 0.1% (v/v) ammonium hydroxide and phase B of 1:1 methanol:acetonitrile with 5 mM ammonium acetate, 0.1% (v/v) ammonium hydroxide. Flow conditions were as follows: 0-1 min, 250 μL/min at 5% B; 1-2 min, 250 μL/min at 5-95% linear gradient of B; 2-2.5 min hold at 95% B at 450 μL/min; 2.5-2.6 min decrease to 5% B at 450 μL/min; 2.6-4.9 min hold at 5% B at 450 μL/min; 4.9-5.0 min return to 250 μL/min at 5% B. With flow at 250 μL/min, ESI ionization was achieved with 25 sheath gas, 5 aux gas; with flow at 450 μL/min, ESI ionization was achieved with 45 sheath gas, 25 aux gas (both N_2_). The polarity was negative for the duration of the run.

Clinical Data Extraction

Clinical outcomes for the first 48 hours following cardiac surgery were investigated. For blood gas measurements, standard clinical ranges noted in patients’ electronic health record were used to determine low, normal, and high values. Post-operative complications included arrhythmias such as junctional ectopic tachycardia and other supraventricular tachyarrhythmias as well as ventricular tachycardia, ventricular fibrillation, premature ventricular contractions, heart block, and bradycardia. We also documented episodes of hypotension, hypertension, blood pressure liability, acute kidney injury, renal insufficiency, low cardiac output syndrome, cardiogenic shock, cardiac arrest, lactic acidosis, respiratory acidosis, and capillary leak. Since many patients had hyperglycemia post-operatively, only patients with hyperglycemia requiring intervention were included. Interventions including extracorporeal membrane oxygenation and transfusion were also recorded. Patients who did not experience the complications and/or interventions noted above were included in the no complications group.

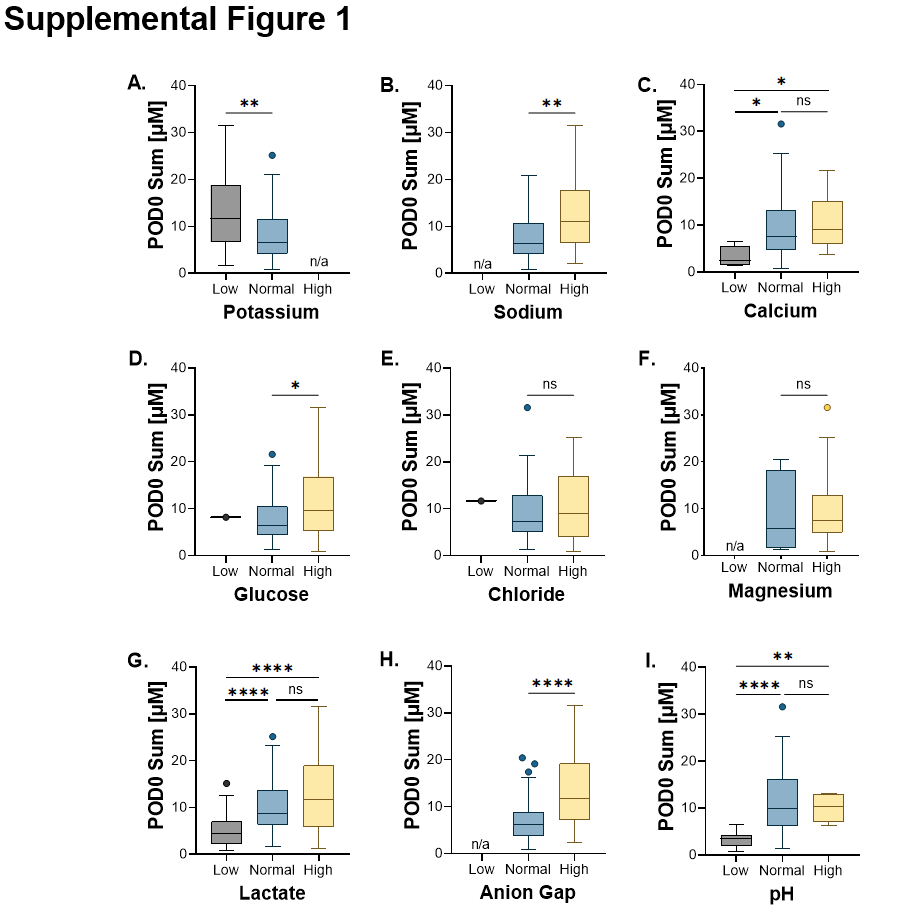
 **Supplemental Data**

**Supplemental Figure 1: Association between post-operative phthalate levels and clinical laboratory values in patients receiving CPB.** Laboratory values are grouped into normal, low, and high ranges according to clinical standards for **A)** potassium, **B)** sodium, **C)** calcium, **D)** glucose, **E)** chloride, **F)** magnesium, **G)** lactate, **H)** ion gap, **I)** pH. n/a indicated when no patients were observed in this group. *Statistical analysis by one-way ANOVA (3 groups) with Welch’s correction for unequal variance, or two-tailed student’s t-test (2 groups). *p<0.05, **p<0.01, ***p<0.005, ****p<0.0001, ns=not significant*

**Supplemental Table 1. Patient demographics according to surgical group.** Values are reported as median [range] or n-value (percent). Inotrope Score and Vasoactive Inotrope Score were calculated at midnight the day of surgery. *4 individual patients died; one patient had multiple independent surgeries over their lifetime and was included in the RBC prime group twice. CPB = cardiopulmonary bypass, LOS = length of stay, Pre-Op = pre-operative, Post-Op = post-operative.

|  | **No CPB** | **Crystalloid Prime** | **RBC Prime** |
| --- | --- | --- | --- |
| **# of Cases** | 11 | 21 | 90 |
| **Age (days)** | 8 [2 - 233] | 1963 [154 - 6324] | 93.5 [1 - 3381] |
| **Weight (kg)** | 3.11 [2.08 - 6.11] | 19.40 [7.60 - 81.80] | 4.38 [2.15 - 29.40] |
| **BSA (m^2^)** | 0.20 [0.14 - 0.32] | 0.79 [0.35 - 1.91] | 0.25 [0.16 - 0.32] |
| **Inpatient (yes)** | 10 (90.9) | 1 (4.8) | 54 (60) |
| **Inpatient Pre-Op LOS (days)** | 7 [2 - 212] | 1 | 9 [1 - 179] |
| **Time to Transfer (days)** | 28.98 [1.13 - 192.8] | 2.01 [0.84 - 13.97] | 5.07 [0.82 - 225.7] |
| **Post-Op LOS (days)** | 44.57 [2.95 - 192.8] | 4.75 [2.83 - 177.0] | 9.97 [2.94 - 235.6] |
| **Mortality Within 30 Days (yes)** | 0 (0) | 0 (0) | n = 4* |
| **Flow Rate (L/min)** | - | 1.89 (0.85 - 4.58) | 0.59 (0.38 - 2.41) |
| **Indexed Flow Rate (L/min/m^2^)** | - | 2.40 [2.38 - 2.43] | 2.41 [2.33 - 2.48] |
| **CPB Duration (min)** | - | 84.0 [42.0 - 257.1] | 113.6 [50.6 - 342.2] |
| **Lowest Venous Temperature on CPB (°C)** | - | 29.40 [25.20 - 32.10] | 26.75 [18.00 - 34.40] |
| **Prime Volume (mL)** | - | 405.8 [195.5 - 790.0] | 113.6 [50.6 - 342.2] |
| **Indexed Prime Volume (mL/m^2^)** | - | 453.1 [283.3 - 878.0] | 951.4 [425.2 - 3435] |
| **Blood Volume in CPB Prime (mL)** | - | - | 100 [50 - 155] |
| **Indexed Blood Volume in CPB Prime (mL/m^2^)** | - | - | 400.0 [107.1 - 833.3] |
| **FFP in CPB Prime (yes)** | 0 (0) | 9 (42.9) | 90 (100) |
| **FFP Volume in CPB Prime (mL)** | - | 100 [75 - 100] | 105 [45 - 150] |
| **Indexed FFP Volume in CPB Prime (mL)** | - | 127 [95.2 - 285.7] | 400 [133.9 - 750.0] |
| **Fluid Balance (mL)** | - | -270 [-942 - 271] | -78 [-778 - 337] |
| **10 mg/kg Methylprednisolone (yes)** | 0 (0) | 0 (0) | 24 (26.7) |
| **Inotrope Score** | 0 [0 - 15] | 0 [0 - 8] | 0 [0 - 15] |
| **Vasoactive Inotropic Score** | 1.425 [0 - 22.25] | 0 [0 - 11] | 5 [0 - 22] |

**Supplemental Table 2. Phthalate chemical exposure according to surgical group.** Values are reported as mean ± standard deviation (SD) with the corresponding range. CPB = cardiopulmonary bypass, DEHP = di(2-ethyhexyl)phthalate, FFP = fresh frozen plasma, MECPP = mono(2-ethyl-5-carboxypentyl)phthalate, MEHHP = mono(2-ethyl-5-hydroxyhexyl)phthalate, MEHP = mono(2-ethylhexyl)phthalate, RBC = red blood cell.

|  | | **DEHP** | | **MEHP** | | **MECPP** | | **MEHHP** | | **Sum** | |
| --- | --- | --- | --- | --- | --- | --- | --- | --- | --- | --- | --- |
|  |  | Mean ± SD | Range | Mean ± SD | Range | Mean ± SD | Range | Mean ± SD | Range | Mean ± SD | Range |
| **No CPB (n=11)** | Pre-Op | 0.939 ± 0.634 | 0.292 - 1.892 | 0.199 ± 0.120 | 0.084 - 0.465 | 0.163 ± 0.085 | 0.032 - 0.324 | 0.068 ± 0.035 | 0.036 - 0.150 | 1.370 ± 0.732 | 0.499 - 2.717 |
|  | POD0 | 0.923 ± 0.279 | 0.390 - 1.284 | 0.179 ± 0.057 | 0.117 - 0.278 | 0.219 ± 0.118 | 0.068 - 0.455 | 0.058 ± 0.024 | 0.025 - 0.094 | 1.381 ± 0.324 | 0.800 - 1.896 |
|  | POD1 | 1.003 ± 0.34 | 0.600 - 1.784 | 0.193 ± 0.097 | 0.110 - 0.446 | 0.237 ± 0.131 | 0.043 - 0.477 | 0.071 ± 0.035 | 0.023 - 0.117 | 1.504 ± 0.393 | 0.886 - 2.195 |
| **Crystalloid prime (n=12)** | Pre-Op | 1.047 ± 0.517 | 0.546 - 2.201 | 0.244 ± 0.197 | 0.095 - 0.747 | 0.086 ± 0.067 | 0.008 - 0.222 | 0.042 ± 0.019 | 0.021 - 0.087 | 1.419 ± 0.646 | 0.772 - 2.795 |
|  | POD0 | 0.988 ± 0.635 | 0.536 - 2.427 | 1.304 ± 1.241 | 0.115 - 3.800 | 0.363 ± 0.309 | 0.040 - 1.111 | 0.096 ± 0.061 | 0.031 - 0.259 | 2.751 ± 1.956 | 0.772 - 6.089 |
|  | POD1 | 0.809 ± 0.174 | 0.542 - 1.174 | 0.220 ± 0.086 | 0.119 - 0.378 | 0.139 ± 0.066 | 0.025 - 0.246 | 0.048 ± 0.028 | 0.017 - 0.099 | 1.216 ± 0.167 | 0.990 - 1.535 |
| **Crystalloid+FFP prime (n=9)** | Pre-Op | 1.216 ± 0.727 | 0.733 - 3.131 | 0.277 ± 0.291 | 0.069 - 1.031 | 0.054 ± 0.014 | 0.033 - 0.073 | 0.038 ± 0.021 | 0.017 - 0.078 | 1.585 ± 1.008 | 1.017 - 4.245 |
|  | POD0 | 1.430 ± 0.545 | 0.803 - 2.497 | 1.585 ± 1.270 | 0.367 - 4.653 | 1.075 ± 0.636 | 0.273 - 2.037 | 0.285 ± 0.204 | 0.049 - 0.729 | 4.375 ± 1.642 | 1.643 - 6.573 |
|  | POD1 | 1.038 ± 0.398 | 0.611 - 1.774 | 0.252 ± 0.052 | 0.172 - 0.312 | 0.219 ± 0.112 | 0.063 - 0.371 | 0.055 ± 0.021 | 0.0303 - 0.085 | 1.563 ± 0.418 | 1.008 - 2.093 |
| **RBC-based prime (n=90)** | Pre-Op | 1.293 ± 1.506 | 0.156 - 11.31 | 0.435 ± 1.317 | 0.057 - 11.73 | 0.142 ± 0.275 | 0.011 - 1.882 | 0.093 ± 0.249 | 0.018 - 2.336 | 1.962 ± 2.558 | 0.346 - 16.92 |
|  | POD0 | 3.790 ± 2.240 | 0.863 - 9.979 | 5.118 ± 4.372 | 0.720 - 20.28 | 1.850 ± 1.463 | 0.221 - 8.092 | 0.644 ± 0.428 | 0.127 - 2.305 | 11.40 ± 6.194 | 2.437 - 31.57 |
|  | POD1 | 1.360 ± 0.8962 | 0.461 - 7.712 | 1.781 ± 3.640 | 0.127 - 26.15 | 1.257 ± 1.185 | 0.035 - 6.545 | 0.442 ± 0.548 | 0.020 - 2.478 | 4.839 ± 4.984 | 0.993 - 33.08 |
